# A multicenter, randomized, double-blind, placebo-controlled ascending dose study to evaluate the safety, tolerability, pharmacokinetics (PK) and pharmacodynamic (PD) effects of Posiphen in subjects with Early Alzheimer’s Disease

**DOI:** 10.1101/2024.03.20.24304638

**Authors:** Douglas Galasko, Martin R. Farlow, Brendan P. Lucey, Lawrence S. Honig, Donald Elbert, Randall Bateman, Jeremiah Momper, Ronald Thomas, Robert A. Rissman, Judy Pa, Vahan Aslanyan, Archana Balasubramanian, Tim West, Maria Maccecchini, Howard H. Feldman

## Abstract

**Background:** Amyloid beta protein (Aβ) is a treatment target in Alzheimer’s Disease (AD). Lowering production of its parent protein, APP, has benefits in preclinical models. Posiphen binds to an iron-responsive element in APP mRNA and decreases translation of APP and Aβ. To augment human data for Posiphen, we evaluated safety, tolerability and pharmacokinetic and pharmacodynamic (PD) effects on Aβ metabolism using Stable Isotope Labeling Kinetic (SILK) analysis.

**Methods:** Double-blind phase 1b randomized ascending dose clinical trial, at five sites, under an IRB-approved protocol. Participants with mild cognitive impairment or mild AD (Early AD) with positive CSF biomarkers were randomized (within each dose arm) to Posiphen or placebo. Pretreatment assessment included lumbar puncture for CSF. Participants took Posiphen or placebo for 21-23 days, then underwent CSF catheter placement, intravenous infusion of ^13^C_6_-leucine, and CSF sampling for 36 hours. Safety and tolerability were assessed through participant reports, EKG and laboratory tests. CSF SILK analysis measured Aβ40, 38 and 42 with immunoprecipitation-mass spectrometry. Baseline and day 21 CSF APP, Aβ and other biomarkers were measured with immunoassays. The Mini-Mental State Exam and ADAS-cog12 were given at baseline and day 21.

**Results:** From June 2017 to December 2021, 19 participants were enrolled, in dose cohorts (6 active: 2 placebo) of 60 mg once/day and 60 mg twice/day; 1 participant was enrolled and completed 60 mg three times/day. 10 active drug and 5 placebo participants completed all study procedures. Posiphen was safe and well-tolerated. 8 participants had headaches related to CSF catheterization; 5 needed blood patches. Prespecified SILK analyses of Fractional Synthesis Rate (FSR) for CSF Aβ40 showed no significant overall or dose-dependent effects of Posiphen vs. placebo. Comprehensive multiparameter modeling of APP kinetics supported dose-dependent lowering of APP production by Posiphen. Cognitive measures and CSF biomarkers did not change significantly from baseline to 21 days in Posiphen vs placebo groups.

**Conclusions:** Posiphen was safe and well-tolerated in Early AD. A multicenter SILK study was feasible. Findings are limited by small sample size but provide additional supportive safety and PK data. Comprehensive modeling of biomarker dynamics using SILK data may reveal subtle drug effects.

**Trial registration:** NCT02925650 on clinicaltrials.gov

## BACKGROUND

Genetic, neuropathological and biomarker evidence support key roles for amyloid beta protein (Aβ) in the pathogenesis of Alzheimer’s Disease (AD) and as a therapeutic target. Aβ is produced by sequential proteolytic cleavages of its parent protein amyloid protein precursor (APP), yielding secreted peptides ending at amino acid residues 37-43 of the Aβ sequence.^1^ Mutations in *PSEN1* or *PSEN2* (which encode proteins that are constituents of the gamma-secretase complex that cleaves APP) result in shifts in APP processing favoring Aβ42.^2–5^ Mutations and rare duplications in the APP gene cause early onset familial AD.^6,7^ Overproduction of APP in Down Syndrome (trisomy 21) results in AD pathology developing inevitably with age.^8^ An APP mutation associated with decreased production of Aβ protects against late onset AD.^9^ Therefore, decreasing APP protein may provide a treatment option for AD. This can be achieved genetically in mouse models, but complete APP knockout is associated with decreased locomotor strength, impaired learning and memory with aging and synaptic changes.^10–12^ Partial reduction of APP is likely to be safer and more practical.

The post-translational regulation of APP includes an iron-responsive element (IRE) in the 5’ untranslated region of APP mRNA. In a screen of compounds that bind to IRE sequences, the (+) and (-) enantiomers of phenserine decreased the translation of APP mRNA, resulting in decreased production of APP and Aβ.^13,14^ While the (-) enantiomer has substantial anticholinesterase activity, the (+) enantiomer (Posiphen, undergoing clinical development as Buntanetap) does not.^15^ Metabolites of Posiphen retain activity in lowering APP translation.^16^ Posiphen lowered APP translation in cell and animal models and showed beneficial effects on cognition in APP and Down syndrome transgenic mice.^17–19^ Because IREs also influence the translation of other proteins, including α-Synuclein, targeting IRE might be beneficial in α-Synuclein disorders such as Parkinson’s Disease (PD) and Lewy Body Dementia (LBD).

A prior phase 1 study of Posiphen determined pharmacokinetic (PK) and pharmacodynamic (PD) data and measured secreted APP (sAPP) in cerebrospinal fluid (CSF).^20^ Posiphen was given orally up to four times per day and had a short plasma half-life. In a Single Ascending Dose study, a daily dose of 160 mg or higher resulted in a high rate of gastro-intestinal side effects. In multiple dose studies, lower doses (20, 40 or 60 mg) given four times per day were safe and well-tolerated. A phase 1 study in healthy adults also included four subjects with Mild Cognitive Impairment (MCI) who underwent serial collection of CSF via lumbar catheter for 12 hours before and after 10 days of dosing with 60 mg four times per day. In this MCI cohort, mean CSF sAPPα, sAPPβ and total tau were decreased at 10 days compared to pretreatment. More recently, Posiphen was given to small cohorts of patients with mild AD (n = 14, 80 mg once per day), and Parkinson’s Disease (10 – 80 mg once per day) for up to 27 days. CSF obtained by lumbar punctures before and after treatment showed a small reduction of APP in the AD study.^21^ There were small changes in CSF Aβ peptide concentrations, with larger decreases for APP biomarkers; however, these differences were not significant. Ascertainment of the relationship between CSF biomarker changes and dose is therefore limited.

To determine pharmacokinetic (PK) and pharmacodynamic (PD) properties of Posiphen and obtain further safety data in the target population of early AD (including MCI and mild AD-dementia), we have conducted a multicenter phase 1b study and evaluated PD effects on Aβ metabolism using Stable Isotope Labeling Kinetic (SILK) analysis in CSF.^22^ SILK enables modelling of production and clearance of CNS proteins based on serial sampling of CSF following a constant infusion of stable isotope labeled leucine.^23^ SILK can identify small changes that can be difficult to determine from longitudinal CSF sampling and has provided insights into how factors such as sleep, circadian rhythms and drugs may affect APP and Aβ metabolism and dynamics.^24–28^

## METHODS

### Overall study design

From June 5, 2017 to December 31, 2021, we conducted the DISCOVER randomized clinical trial at five participating sites, coordinated by the Alzheimer’s Disease Cooperative Study (ADCS) at UCSD, with funding from the National Institute on Aging and investigational product provided by Annovis, Inc. This study was approved by each participating site’s IRB and was registered on https://www.clinicaltrials.gov/ as NCT02925650. The trial was conducted according to the guidelines of good clinical practice (GCP) and was monitored by the ADCS. A Data Safety Monitoring Board (DSMB) reviewed data for each cohort and for the study overall. The study was designed as a sequential cohort ascending dose trial, with subjects receiving 21-23 days of Posiphen vs placebo. Within each study cohort, participants were randomly assigned to receive either Posiphen or placebo for 21-23 days, followed by a confinement visit, in which they were admitted to an inpatient observation unit, had a lumbar intrathecal catheter and intravenous line placed, followed by intravenous infusion of ^13^C_6_-leucine and serial sampling of blood and CSF over 36 hours for Stable Isotope Labeling and Kinetics (SILK).

There were three primary objectives of the study: 1) to assess safety and tolerability of Posiphen in people with early AD, including people with Mild Cognitive Impairment or mild dementia due to Alzheimer’s Disease, over a dose range of 60 mg given once, twice or three times per day; 2) to determine PK for Posiphen and its N1 and N8 metabolites by measuring their levels in CSF and plasma at multiple time points during the first 24 hours of CSF catheterization; 3) to determine PD by using SILK data to model the fractional synthesis rate (FSR) of Aβ40, the most abundant form of secreted Aβ, and also to assess changes in CSF levels of Aβ40 from pretreatment to 21 days post-treatment. Additional objectives included: establishing the feasibility of carrying out a multi-center SILK study; assessing pharmacodynamic effects of Posiphen on other biomarkers in CSF including Amyloid Beta 38 (Aβ38), Aβ40, Aβ42, soluble Amyloid-β Precursor Protein alpha (sAPPα), soluble Amyloid-β Precursor Protein alpha (sAPPβ) and t-Tau; and evaluating cognitive and/or neuropsychiatric effects of Posiphen. Because there have been substantial advances in methods of analysis of SILK data since this study was originally designed, we also carried out post-hoc advanced modeling of these data to determine APP synthesis rates. Further details of the study, including a detailed protocol, are available at https://www.clinicaltrials.gov/ under NCT02925650.

### Study drug and ^13^C_6_-leucine

Posiphen (investigational new drug #72 654) was manufactured according to Good Manufacturing Practice (GMP) regulations. It was packaged as gelatin capsules, without excipients or fillers and matching placebo capsules (non-lactose compound) which were prepared with an inert inactive excipient generally recognized as safe for human pharmaceutical use by Frontage Laboratories, Inc. Capsules were packaged in identical blister packs and provided by Frontage to study sides to preserve the blind.

^13^C_6_-leucine was supplied by C2N, Inc, (St Louis, MO) as a powder, manufactured under MPT grade specifications. Some of the site investigational pharmacies were unable to prepare ^13^C_6_-leucine by following the prior Washington University/C2N protocol, and in September 2019, GMP-grade ^13^C_6_-leucine was obtained from C2N, prepared by PCI Study Services, Inc, (San Diego, CA), and shipped as vials to sites within the week prior to the confinement visit. Sites followed a protocol by diluting the ^13^C_6_-leucine in sterile normal saline, with dosing dependent on body weight, and intravenous infusion with an initial bolus of 3 mg/kg followed by steady infusion over the remainder of 9 hours at 2 mg/kg/h.

### Participants

Study participants were included if they met the following criteria: 1) men or women aged 55-89; 2) female participants were postmenopausal or not of childbearing potential, 3) female participants test negative with a urine pregnancy test at the screening visit); 4) MCI-AD (NIA-AA criteria)^29^ or mild AD with Mini-Mental State Examination (MMSE) 17-30 and global Clinical Dementia Rating (CDR)^30^ 0.5 – 1.0; participant likely to tolerate all study procedures as judged by the local site PI: 5) CSF biomarkers consistent with AD: Aβ42/Aβ40 < 0.131 assayed by mass spectrometry (C2N Diagnostics, St Louis, MO – unpublished data for the CSF cutoff for amyloid positivity was provided by C2N); 6) general cognitive abilities sufficient for the participant to provide informed consent; 7) study partner available; 8) no evidence of current suicidal ideation or prior suicide attempt; 9) MRI in the past 12 months without evidence of infection, infarction, or other focal lesions; 10) stable doses of medications for 4 weeks and of AD medications (acetylcholinesterase inhibitors and/or memantine) for 12 weeks prior to screen; 11) adequate vision hearing and good general health; 12) fluent English speaker.

Exclusion criteria were: 1) major psychiatric disorder; 2) other neurodegenerative disease; 3) dementia other than AD; 4) seizure disorder; 5) clinically significant abnormalities on screening laboratory studies or EKG results; 6) current serious or unstable illness; 7) four or more microinfarcts on brain MRI; 8) cancer currently or in past 3 years (other than skin or stable untreated prostate cancer); 9) alcohol abuse or dependence or drug abuse; 10) participation in another clinical trial and have taken active investigational product within 16 weeks before screening; 11) resides in skilled nursing facility; 12) contraindications to lumbar puncture and catheterization, e.g., infection in skin close to site; major orthopedic abnormality, disorder of coagulation or use of anticoagulant drug; 13) deep brain stimulator present.

Participants who were recruited at each participating site signed an informed consent document, and eligibility was determined during a screening period of up to 42 days. This allowed adequate time for the analysis of CSF to determine amyloid status.

### Randomization

Participants who signed an informed consent and met screening eligibility requirements were randomly assigned to receive 60 mg Posiphen once daily or placebo (with an allocation of 5 to Posiphen, 3 to placebo) by a stratified, random permuted blocked treatment assignment method, stratified by site. Following dosing of the initial 8 participants and DSMB review of safety and tolerability, the next dose group was 60 mg of Posiphen or placebo twice daily (BID) (randomly assigned as 5 to Posiphen, 3 to placebo). Following the dosing of this group of 8 participants and DSMB review of safety and tolerability, the next dose group was 60 mg of Posiphen or placebo three times daily (TID) (randomly assigned as 5 to Posiphen, 3 to placebo).

### Study visits

The screening visit included history, examination, review of medical records and medications, vital signs, physical and neurological examination, cognitive testing with the MMSE and Logical Memory test (Wechsler Memory Scale-revised), EKG, blood draw for routine laboratory tests, lumbar puncture and if needed, brain MRI. If eligible, participants underwent a baseline visit, with cognitive assessment using the MMSE and ADAS-cog12, behavioral assessments, Research Satisfaction Survey, safety assessments, and review of concurrent medications and adverse events, and they received a supply of the study drug (or placebo), which they started the day after the baseline visit. A telephone assessment was done at day 14 after baseline, to assess adverse events, concomitant medication changes and remind participants/study partners about the confinement visit. A pre-confinement visit took place 1-3 days before the confinement visit, following treatment for 20 days, and included physical and neurological examination, cognitive and behavioral assessments (see below), safety assessments (including clinical safety labs), and a review of compliance, concurrent medications and adverse events. The study drug or placebo was continued until the day before the confinement visit.

For the confinement visit, subjects arrived in the early morning after an overnight fast. Physical and neurological examination, vital signs, and assessment of compliance were completed and blood was drawn for safety laboratory tests. An intravenous line and a second line for repeated blood draw were placed. To sample CSF, the intrathecal space was accessed using a Perifix 18G Touhy Epidural Needle (90mm), and a Polyamide Epidural Catheter (20G Closed Tip) was inserted and fixed in place by an Anesthesiologist. Participants received the last day’s dosing of study drug or placebo. Initial blood and CSF samples were drawn, intravenous infusion of ^13^C_6_-leucine was started and continued for 9 hours, during which time subjects received a low leucine diet. Blood and CSF were sampled every 2 hours for 36 hours. After 36 hours, intravenous lines and the CSF catheter were removed, vital signs were measured, physical exam performed, and blood and CSF were sent for routine laboratory studies. Subjects were observed for 12 hours after removal of the CSF catheter. If postural headache was present, a blood patch was performed. Once stable, participants underwent a brief exam and vital sign check and were discharged. They received a phone call 24 hours after discharge to check on their wellbeing.

### Outcome measures

Safety measures included adverse events, vital signs, EKG and laboratory tests. The Mini-Mental State Examination (MMSE)^31^, Alzheimer’s Disease Assessment Scale-cognitive (ADAS-cog12)^32^ and Neuropsychiatric Inventory (NPI)^33^ were used as cognitive and behavioral outcomes,.

For biochemical analysis to derive data for SILK, ^13^C_6_-leucine was analyzed in plasma and CSF by mass spectrometry, and Aβ peptides in CSF were analyzed by immunoprecipitation-mass spectrometry (IP-MS) as previously described,^23^ by C2N Diagnostics (St Louis, MO). CSF levels of sAPPα, sAPPβ, Aβ38, 40 and 42 were analyzed by the ADCS Biomarker Core using Mesoscale Discovery (MSD) electrochemiluminescent assays (Mesoscale Discovery, Rockville, MD). Levels of Posiphen and its N1 and N8 metabolites in plasma and CSF were measured by Charles River Laboratories (Montreal, CA) by high-performance liquid chromatographic mass spectrometry, with use of deuterated Posiphen (procured by Charles River Laboratories) as an internal standard.

### Statistical analysis

The study was powered based on SILK data on Fractional Synthesis Rate (FSR)^23^ for Aβ40, measured in people with AD, using prior data from patients with mild AD (provided by C2N Diagnostics), and was modeled to evaluate a primary analytic goal of demonstrating a monotonic dose response relationship to Posiphen treatment. Based on a two-sided t-test for the difference between two independent means at a significance level (alpha) of 0.05 and a statistical power of 80%, a sample size of 5 subjects in each treatment group (n active = 15) and 9 controls would provide at least 80% statistical power to detect a 27% change in the mean Aβ40 FSR. For comparison, the rare APP A673T genetic variant that protects against late-onset AD is associated with 9-26% lower sAPPβ and Aβ42 levels in CSF compared to age-matched conrols.^34^

A prespecified Statistical Analysis Plan (SAP) was prepared and finalized before the database lock occurred. For safety and tolerability, frequencies of adverse events, serious adverse events and laboratory abnormalities between the participants across the three groups are compared, using Pearson’s χ^2^ or ANOVA, as appropriate. Plasma concentration-time data are analyzed by non-compartmental methods using WinNonlin version 6.2.1 or greater. Calculations are based on the actual sampling times recorded during the study. For PK analysis, only the subjects receiving Posiphen are included. From the plasma concentration-time data, the following PK parameters were estimated, as data permit: area under the concentration-time curve from 0 to 24 hours (AUC_0-24_), maximum observed plasma concentration (C_max_,), time of maximum observed plasma concentration (T_max_), terminal elimination half-life (t_1/2_,), apparent volume of distribution (V/F), and apparent oral clearance (Cl/F).

For SILK, the fraction of labeled Aβ for Aβ38, 40, and 42 during 36 hours after administration of study drug for each subject was calculated, as described. The FSR measures were calculated by fitting a line to the upslope of the CSF Aβ labeling data. It was assumed that the upslope occurred between hours 6-16 h after the start of confinement. FSR for treated and untreated subjects were compared by analysis of variance (ANOVA). The percentage of newly synthesized and degraded Aβ was estimated over 36 hours by multiplying the plasma concentration of Aβ measured at the start of confinement (i.e. at catheter placement; time = 0) by the percentage of Aβ that contains ^13^C_6_-leucine at each time point. This can be used to estimate the AUC of the drug for decreasing the amount of newly produced Aβ.^24^ As a post hoc analysis, the FSR measurement was also performed with different assumptions: the timeframe for measuring the upslope was automatically detected based on the slope of the SILK curve (see supplementary materials for method and code). As an additional post hoc analysis, a newer model for SILK that take into count the multiple compartments and CSF and other spaces involved in APP metabolism^35^ was also used to estimate APP production rates, as described below.

Changes in CSF biomarkers from the screening to confinement (sample at time of placement of catheter) were compared in the treatment and placebo groups by ANOVA, with models that included age, sex and baseline MMSE, and further analysis stratified by treatment dose. A similar approach was taken to analyze changes in MMSE, ADAS-cog and NPI. Data were analyzed using R (version 4.2.2).

### Detailed modeling of APP metabolism

APP and Aβ production and clearance rates were determined using a recently described comprehensive model.^35^ The model accounts for the bi-hourly withdrawal of 6 mL of CSF, which is known to alter the lumbar CSF concentration of Aβ.^25^ The model estimates rates of APP production, enzymatic production of Aβ, transport of Aβ from interstitial fluid (ISF) to CSF, enzymatic degradation of Aβ, blood-brain barrier transport of Aβ, as well as CSF mixing throughout the subarachnoid space (SAS) and clearance of CSF from cranial and spinal subarachnoid space (SAS). For the DISCOVER trial, MRI volumetric scans were not available, so CSF volumes (cranial, cisternal and ventricular) were estimated based on the subject’s age using data from the previous study (see Supplementary Materials). The model fits the SILK and lumbar concentration data by varying the following four adjustable parameters: *k*_BPD40_ (rate of clearance of Aβ_40_ from ISF), SF_40_ (scaling factor for Aβ_40_to account for mass spec calibration errors), *Q*_CSF_ (rate of CSF production and clearance) and *Q*_osc_ (parameter describing magnitude of CSF mixing between SAS compartments). In addition to the adjustable parameters, three additional parameters were systematically varied: *f*_VCSF_ (accounting for errors in the estimate of CSF volumes), *V*_SP_ (accounting for errors in the estimate of volume of spinal CSF) and *Q*_leak_ (accounting for potential, but undocumented, CSF leaks around the indwelling lumber catheter).

Additionally, simulations were repeated limiting *k*_BPD40_ to a range of smaller values (0.05-0.2 h^-1^) and a range of larger values (0.2-0.35 h^-1^), based on the observation of local minima in both regimes. In total, 128 parameter optimizations per subject were performed, determining the adjustable parameters by fitting the SILK data while holding the systematically varied parameters at combinations of the following values: *f*_VCSF_ = 0.5, 0.75, 1.0, 1.25; *V*_SP_ = 40, 60, 80 and 100 mL; *Q*_leak_ = 0, 5, 10, 15 mL/h. Then, the parameter combinations were rank-ordered based on the goodness-of-fit to both the SILK and early Aβ lumbar concentration data (0-15 h). The Aβ production rates output from the model were averaged for all scans with sum-of-square residual fits within 20% of the best value for both SILK data and Aβ lumbar concentration data. Later Aβ lumbar concentration data were not considered due to substantial variation in the effect of sleep on CSF Aβ (sleep was not controlled in this study and most experiments began between 9-11 am local time). CSF Aβ concentrations prior to 11 am were also excluded due to the effects of sleep the previous night.^35^

## RESULTS

The trial enrolled 19 of the pre-planned 24 participants, after being significantly impacted by several substantial delays, including logistical problems with the ^13^C_6_-leucine supply and extended shut down periods of inpatient research units during the Covid-19 pandemic between March 2020 and December 2021, when the trial ended. The 60 mg once per day and twice per day dose cohorts were successfully enrolled and completed. The 60 mg three times per day group enrolled and completed only one participant. The disposition of participants is shown in Figure 1.

**Figure 1.**
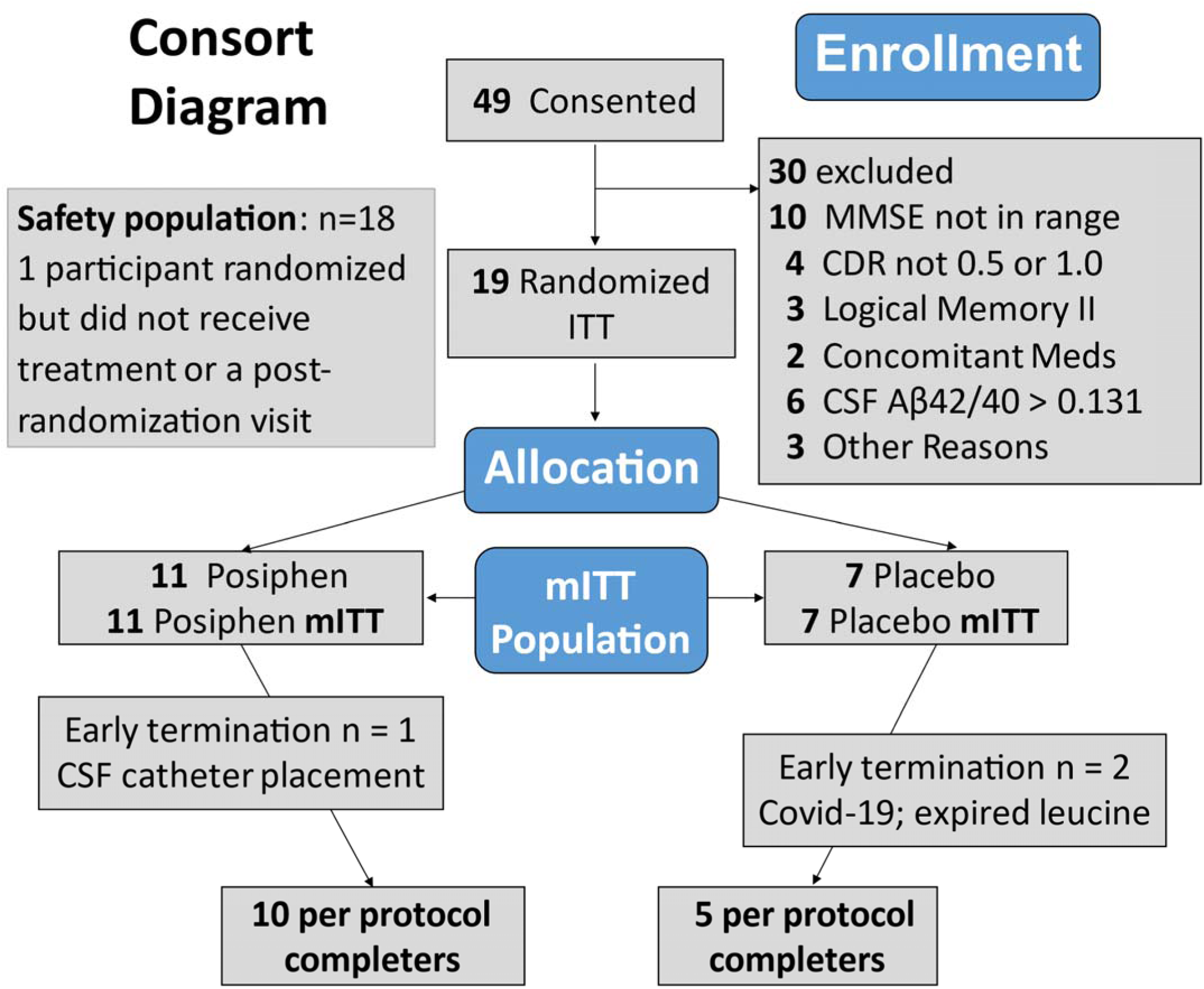
CONSORT diagram.

Forty-nine people were screened, and nineteen were enrolled. One participant was randomized but not enrolled because of a research center shut down due to Covid-19, two participants did not have a confinement visit due to changing regulations about research during the Covid-19 pandemic, and in one participant, the CSF catheter could not be placed to allow successful repeated sampling of CSF. The study procedures were completed in 5 control and 10 active drug treatment participants. Demographic, clinical data and concomitant medications relevant for AD and mood are shown in Table 1 for the nineteen trial participants who were randomized.

**Table 1.**
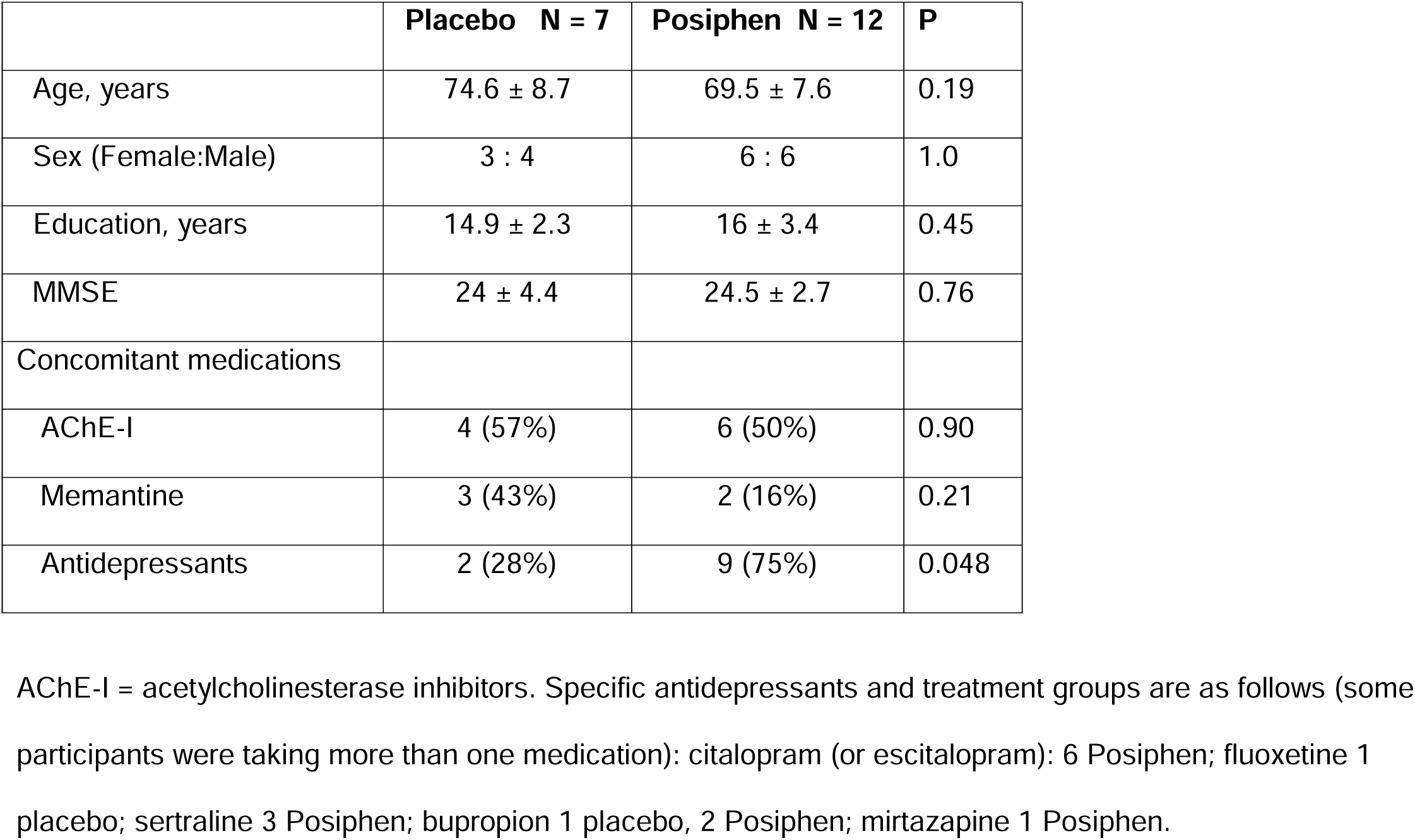
Characteristics of randomized participants.

|  | **Placebo N = 7** | **Posiphen N = 12** | **P** |
| --- | --- | --- | --- |
| Age, years | 74.6 ± 8.7 | 69.5 ± 7.6 | 0.19 |
| Sex (Female:Male) | 3 : 4 | 6 : 6 | 1.0 |
| Education, years | 14.9 ± 2.3 | 16 ± 3.4 | 0.45 |
| MMSE | 24 ± 4.4 | 24.5 ± 2.7 | 0.76 |
| Concomitant medications |  |  |  |
| AChE-I | 4 (57%) | 6 (50%) | 0.90 |
| Memantine | 3 (43%) | 2 (16%) | 0.21 |
| Antidepressants | 2 (28%) | 9 (75%) | 0.048 |

AChE-I = acetylcholinesterase inhibitors. Specific antidepressants and treatment groups are as follows (some participants were taking more than one medication): citalopram (or escitalopram): 6 Posiphen; fluoxetine 1 placebo; sertraline 3 Posiphen; bupropion 1 placebo, 2 Posiphen; mirtazapine 1 Posiphen.

### Safety and Tolerability

Posiphen was safe and was generally well tolerated among the 18 participants who contributed to the safety analyses (7 placebo and 11 who received Posiphen). Vital signs showed no significant changes.

Adverse events (AEs) did not differ in frequency between the groups and there was no pattern of organ system-related AEs. The only Serious Adverse Events in the Posiphen or placebo group were headaches in relation to CSF catheterization, reported in 8/15 participants, 4 while the CSF catheter was in situ; 3 after removal; and 1 at both times. Five participants needed blood patches, and in all 5 the headache rapidly resolved.

ECG parameters showed no significant changes or differences between Posiphen and placebo groups. Laboratory safety parameters showed no significant differences in the placebo and treatment groups and there were no clinically significant changes associated with Posiphen.

### Pharmacokinetics

Data for Posiphen plasma and CSF analysis are shown in Figure 2.

**Figure 2.**
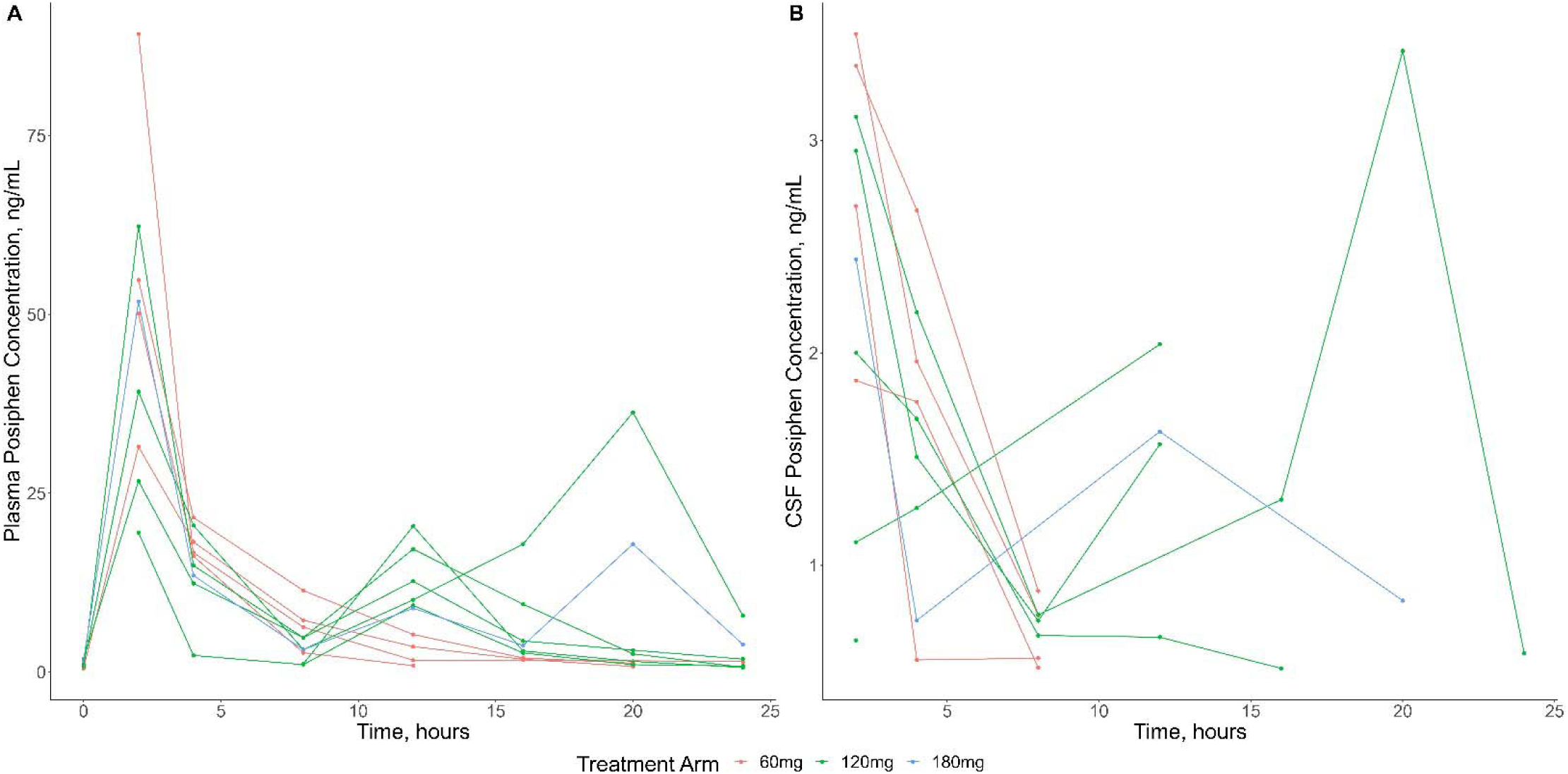
Pharmacokinetics of Posiphen in A) plasma and B) CSF.

Noncompartmental pharmacokinetic parameters were estimated using Phoenix WinNonlin v8.3 (Certara, Princeton, NJ). Levels and parameters (Cmax, AUC) of posiphen, n-1 and n-8 metabolites were highly correlated in plasma and CSF. For the BID and TID doses the expected post dose plasma peaks appeared attenuated.

Pharmacokinetic parameters of Posiphen and its N1 and N8 metabolites in plasma and CSF are shown in Table 2 (mean drug levels over 24 hours; in ng/mL), and additional parameters in Tables 3 and 4. Concentrations of Posiphen and its metabolites were not detectable in any plasma or CSF samples from participants in the placebo arm. Posiphen plasma exposure did not increase proportionally with more frequent dosing. N1 and N8 AUC_0-24_ in plasma increased approximately proportionally between once daily and twice daily dosing. CSF concentrations of Posiphen, N1, and N8 metabolites were measurable in the majority of participants. Concentrations of Posiphen, N1, and N8 were positively correlated in plasma and CSF (e.g., Spearman’s R ranged from 0.73 – 0.82 for AUCs in CSF).

**Table 2.**
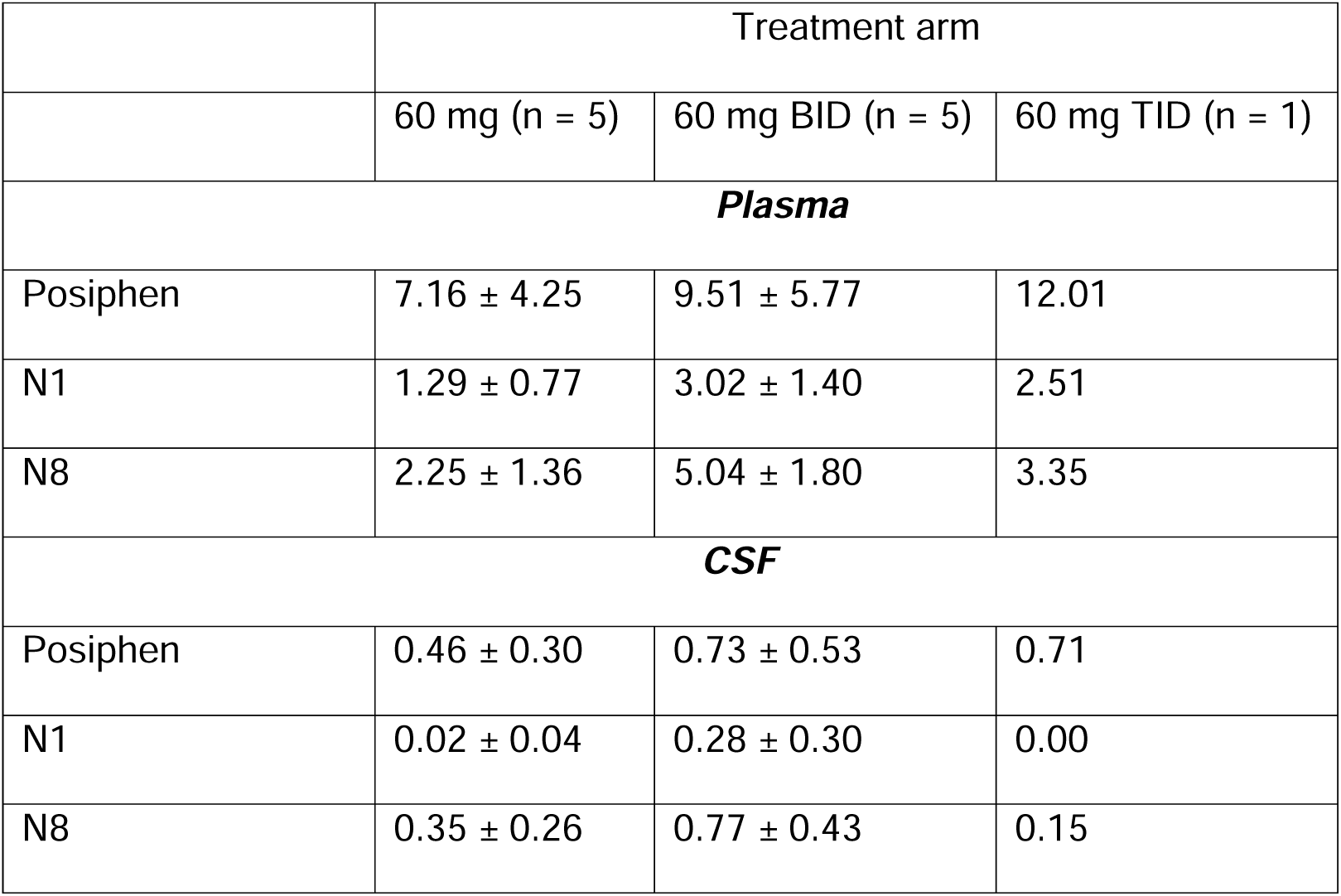
Pharmacokinetic data: mean levels of Posiphen (ng/mL) averaged over 24 hours of CSF sampling.

|  | Treatment arm |  |  |
| --- | --- | --- | --- |
|  | 60 mg (n = 5) | 60 mg BID (n = 5) | 60 mg TID (n = 1) |
| **Plasma** |  |  |  |
| Posiphen | 7.16 ± 4.25 | 9.51 ± 5.77 | 12.01 |
| N1 | 1.29 ± 0.77 | 3.02 ± 1.40 | 2.51 |
| N8 | 2.25 ± 1.36 | 5.04 ± 1.80 | 3.35 |
| **CSF** |  |  |  |
| Posiphen | 0.46 ± 0.30 | 0.73 ± 0.53 | 0.71 |
| N1 | 0.02 ± 0.04 | 0.28 ± 0.30 | 0.00 |
| N8 | 0.35 ± 0.26 | 0.77 ± 0.43 | 0.15 |

**Table 3.** Pharmacokinetic parameters for Posiphen and metabolites in plasma.

|  | Treatment arm |  |  |
| --- | --- | --- | --- |
|  | 60 mg (n = 4) | 60 mg BID (n = 5) | 60 mg TID (n = 1) |
| **Posiphen** |  |  |  |
| AUC<sub>0-24</sub> (hr*ng/mL) | 212.32 (18.14) | 195.80 (69.26) | 288.25 |
| C<sub>max</sub> (ng/mL) | 52.70 (44.67) | 30.39 (51.86) | 51.80 |
| T<sub>max</sub> (hr) | 2 (2 – 2) | 2 (2 – 12) | 2 |
| t<sub>1/2</sub> (hr) | 3.55 (48.65) | . | . |
| V/F (L) | 1403.1 (59.23) | . | . |
| Cl/F (L/hr) | 274.04 (17.50) | . | . |
| **N1 metabolite** |  |  |  |
| AUC<sub>0-24</sub> (hr*ng/mL) | 36.60 (19.24) | 65.06 (58.61) | 60.18 |
| C<sub>max</sub> (ng/mL) | 5.43 (21.98) | 6.96 (32.01) | 5.48 |
| T<sub>max</sub> (hr) | 2 (2 – 4) | 12 (2 – 20) | 2 |
| **N8 metabolite** |  |  |  |
| AUC<sub>0-24</sub> (hr*ng/mL) | 66.02 (20.83) | 113.65 (44.01) | 80.40 |
| C<sub>max</sub> (ng/mL) | 6.81 (25.10) | 10.01 (27.79) | 5.41 |
| T<sub>max</sub> (hr) | 3 (3 – 4) | 12 (2 – 20) | 2 |

AUC<sub>0-24</sub>, area under the concentration-time curve from 0 to 24 hours; C<sub>max</sub>, maximum observed concentration; T<sub>max</sub>, time of maximum observed concentration; t<sub>1/2</sub>, terminal elimination half-life; V/F, apparent volume of distribution; Cl/F, apparent oral clearance. Data are presented as geometric mean (% geometric coefficient of variation) except T<sub>max</sub> which is presented as *median* (min-max). t<sub>1/2</sub>, V/F, and Cl/F are reported only for the 60 mg daily group.

**Table 4.**
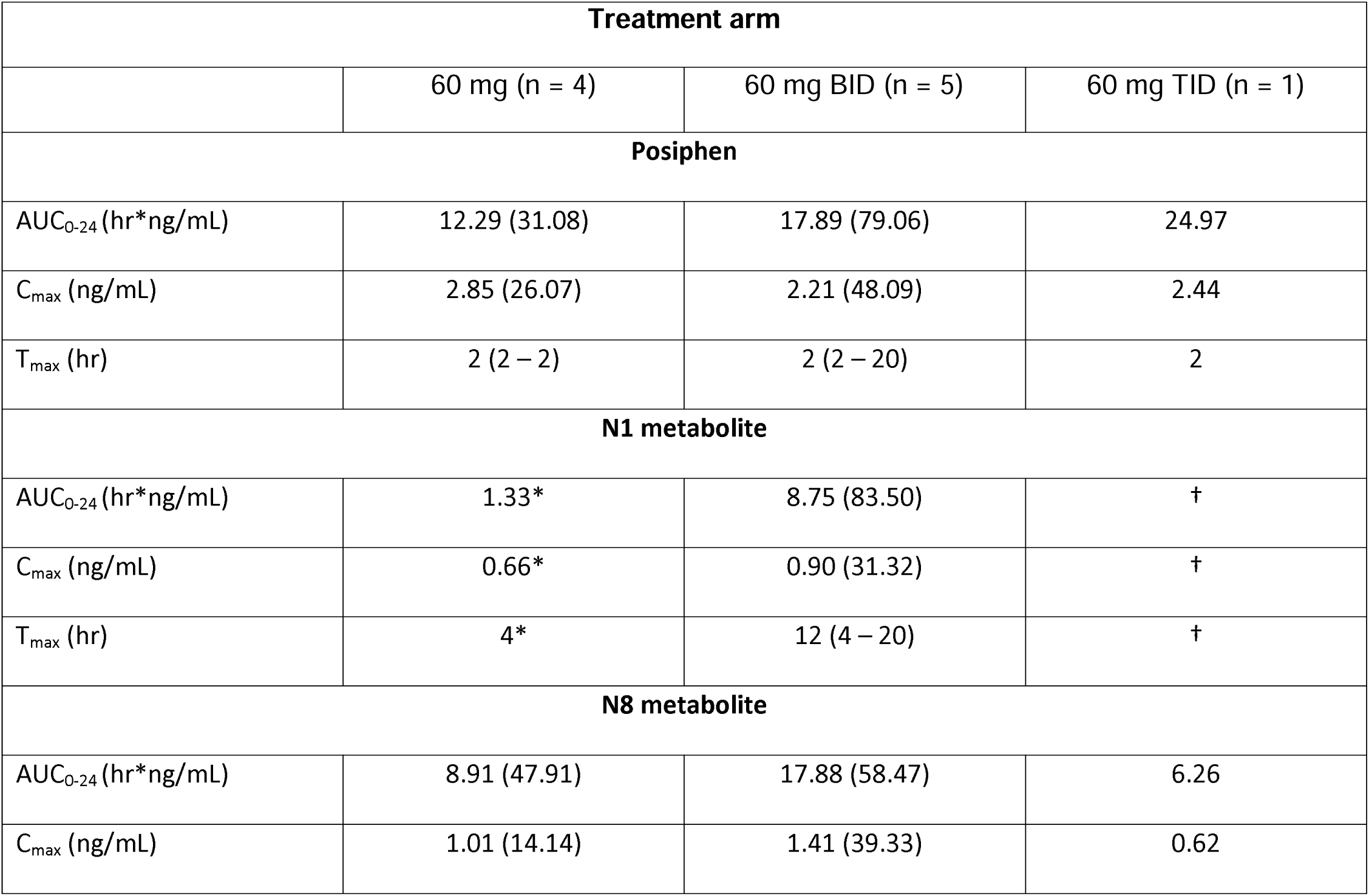
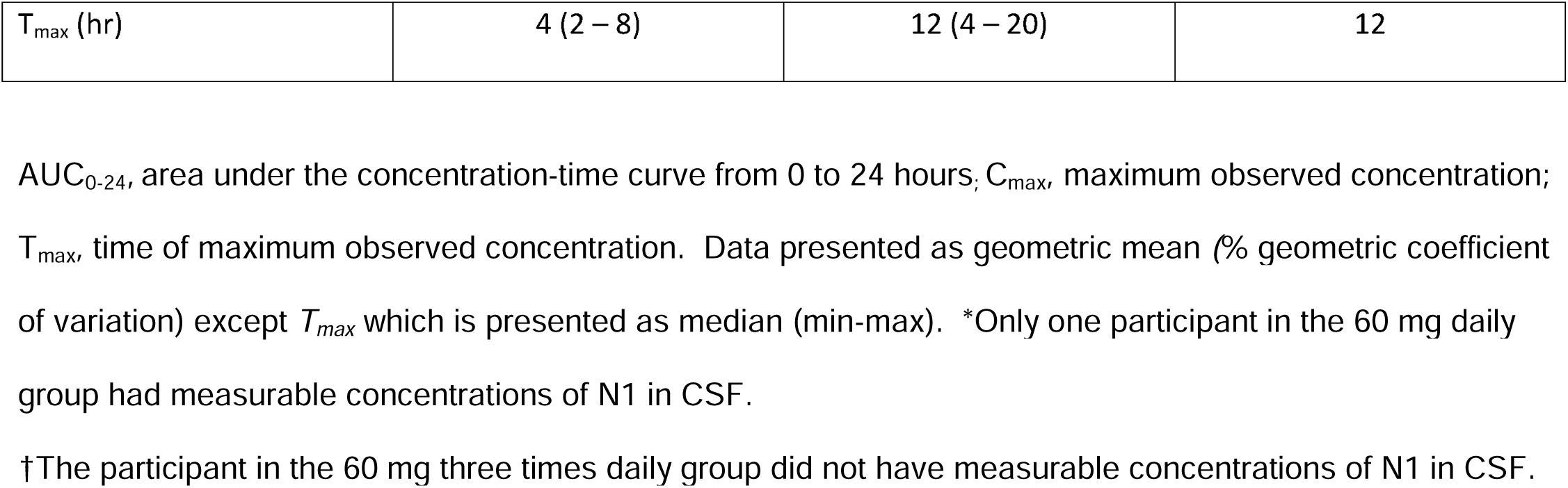
Pharmacokinetic parameters for Posiphen and metabolites in CSF.

### Pharmacodynamics

The SILK time courses for all subjects are shown in Figure 3. The prespecified analyses of FSR for CSF Aβ40 after 21 days of continuous treatment, that modeled FSR from hours 6-16 of the CSF catheter sampling data and assumed linearity during this period, were supplemented with an automated method for calculating the FSR. In both of these analytical approaches, we found no significant effect of Posiphen vs. placebo on FSR for CSF Aβ40, and no dose-dependent effect (Figure 4). The one participant who completed the 180 mg dose had a lower FSR (0.018) than other participants in our study (range 0.298 – 0.462).

**Figure 3.**
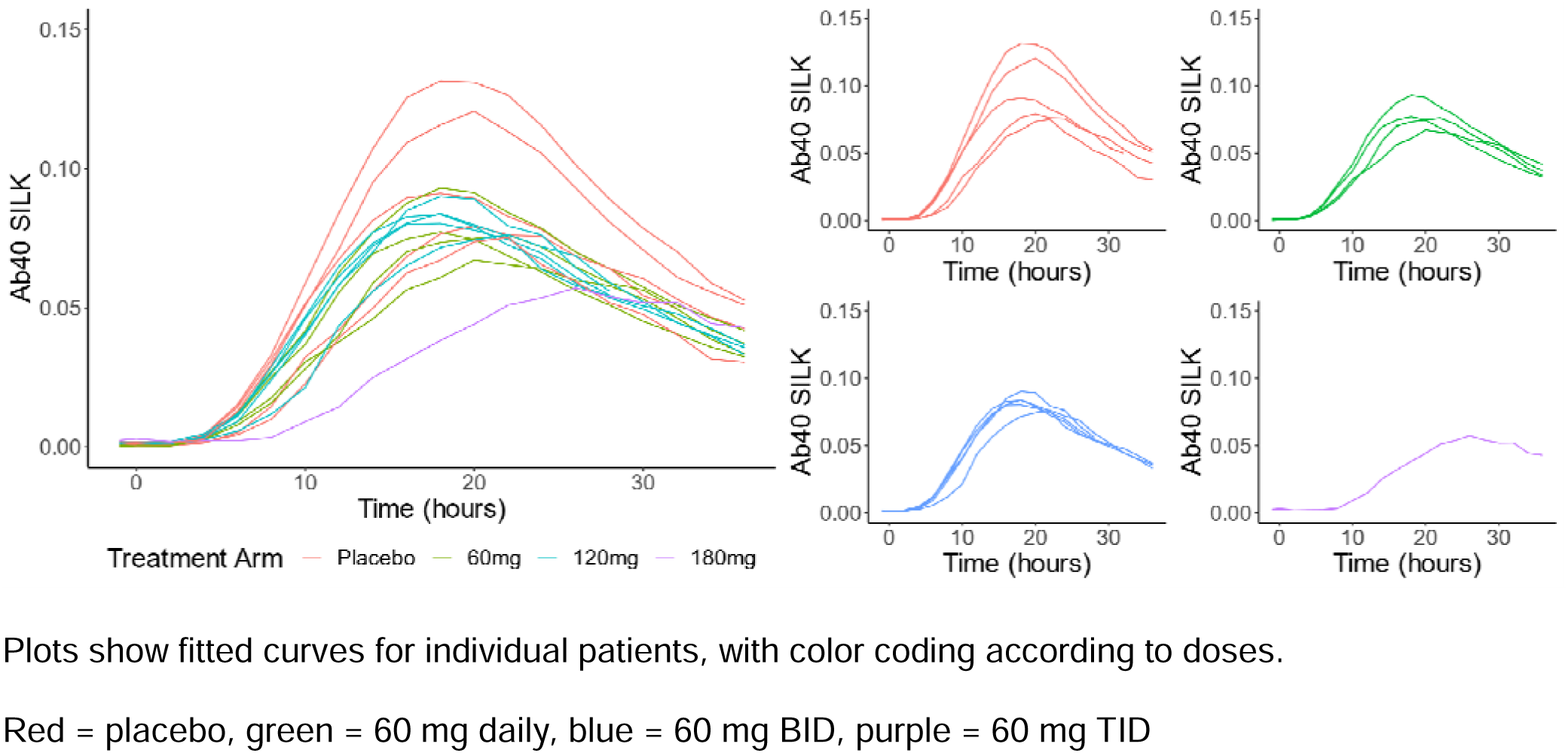
Time course of CSF catheter sampling and SILK data for Aβ40.

**Figure 4.**
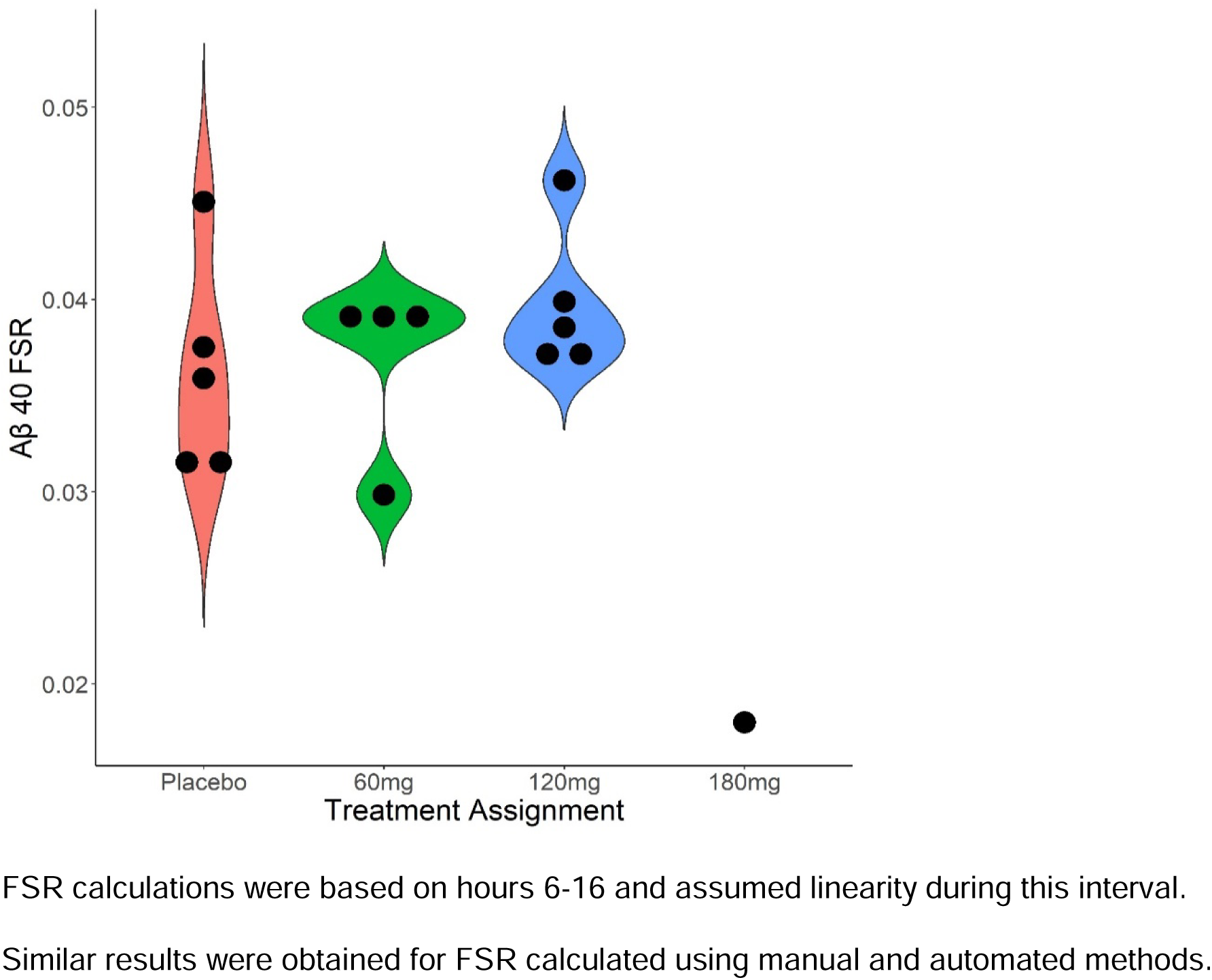
Dose group data for Fractional Synthesis rates for Aβ40.

In a previous observational study with 38 amyloid positive subjects, the mean FSR for CSF Aβ40 was 0.0361 +/- 0.00954 (SD), comparable to the results here. FSR data for CSF Aβ38 and Aβ42 were also calculated. There was no significant effect of Posiphen vs placebo on FSR estimates for Aβ38 and Aβ42. FSR estimates for CSF Aβ38 and CSF Aβ40 were highly correlated (r = 0.99 (p<0.001)). The correlation between FSRs for Aβ40 and 42 was lower (r = -0.42 (p=0.12)), which likely reflects the effects of plaques and insoluble Aβ in the brain on specifically modifying Aβ42 kinetics.^22^ Single timepoint measures of CSF levels of sAPPα, sAPPβ, total tau, Aβ38, 40 and 42 were compared from CSF at the screening lumbar puncture to CSF sampled when the lumbar catheter was inserted at the confinement visit (Table 5). There were no significant differences between Posiphen overall vs placebo, and no dose-dependent differences. The direction of change for Posiphen treatment was to decrease levels of Aβ38, 40 and 42 (after controlling for the slight increase in levels found in the placebo group), and to increase sAPPα, sAPPβ and total tau, but the magnitude of change was very small.

**Table 5.** Longitudinal changes in CSF biomarkers from screening to start of confinement visit.

|  | Arm | Screening | Day 21 | LS mean change | SEM | Treatment difference (Posiphen – placebo)<sup>a</sup> |
| --- | --- | --- | --- | --- | --- | --- |
| **Aβ38**<br>n = 17 | Placebo | 2.90 ± 1.02 (n=7) | 3.52 ± 1.21 (n=7) | 0.593 | 0.199 | -0.202 (0.27) |
| | Posiphen | 2.58 ± 0.85 (n=10) | 2.94 ± 0.89 (n=9) | 0.391 | 0.173 | p = 0.47 |
| **Aβ40**<br>n=17 | Placebo | 14.2 ± 5.3 (n=7) | 17.4 ± 6.14 (n=7) | 3.13 | 1.151 | -1.4 (1.57) |
| | Posiphen | 12.7 ± 4.46 (n=10) | 14.5 ± 4.34 (n=9) | 1.73 | 0.998 | p= 0.39 |
| **Aβ42**<br>n = 17 | Placebo | 1.02 ± 0.38 (n=7) | 1.24 ± 0.39 (n=7) | 0.211 | 0.0753 | -0.104 (0.102) |
| | Posiphen | 0.89 ± 0.24 (n=10) | 0.99 ± 0.24 (n=9) | 0.107 | 0.0653 | p = 0.33 |
| **sAPPα**<br>n = 18 | Placebo | 497 ± 39.6 (n=7) | 497 ± 21.91 (n=4) | **-3.50** | 9.07 | 9.99 (10.8) |
| | Posiphen | 476 ± 90.2 (n=11) | 483 ± 99.5 (n=10) | 6.49 | 5.75 | p = 0.38 |
| **sAPPβ**<br>n = 18 | Placebo | 664 ± 167 (n=7) | 669 ± 130 (n=4) | **-28.6** | 26.5 | 47.9 (31.6) |
| | Posiphen | 591 ± 225 (n=11) | 608 ± 243 (n=10) | 19.1 | 16.8 | p = 0.16 |
| **Total tau**<br>n = 18 | Placebo | 857 ± 262 (n=7) | 821 ± 278 (n=4) | **53.7** | 124.9 | 133 (149) |
| | Posiphen | 751 ± 404 (n=11) | 849 ± 369 (n=10) | **186.4** | 79.1 | p = 0.39 |

CSF biomarker concentrations are expressed as pg/mL.
LS = least squares. SEM = standard error of mean
<sup>a</sup> Analyses of changes in CSF biomarkers used prespecified ANOVA models, with age and sex as covariates.

### Exploratory pharmacodynamic modeling

Modeling of FSR was developed over 15 years ago when the SILK methods were first developed, studied and reported, and fractional synthesis rate (FSR) and fractional clearance rate (FCR) parameters were modeled. Subsequently, there have been more detailed models of production and disposition of APP and its metabolites in the brain and CSF, which we were able to apply to estimate APP production rates, described above.^35^ The production rates across 128 optimizations tended to be tightly clustered within a subject. However, some parameter combinations led to reasonable fits by driving *Q*_osc_ towards zero, leading to predicted ratios of ISF:lumbar Aβ ratios > 10 and extremely high values of Aβ production rates. This was avoided by penalizing ISF:lumbar Aβ ratios > 10 during optimization, as ratios >3-4 are not supported by microdialysis studies.^36^

The APP production rates trended lower in subjects receiving 120 or 180 mg of Posiphen per day. However, the differences were not significant by non-parametric analysis (Figure 5A). Additionally, the lumbar CSF concentrations prior to drug exposure trended lower in the 120 mg and 180 mg groups (Figure 5B), suggesting the possibility that these subjects had lower APP production rates prior to drug exposure due to natural variation. Furthermore, the number of subjects per group was less than the study design due mainly to COVID restrictions. With these caveats in mind, a model that included the pre-drug lumbar CSF concentration as a covariate showed significance between the placebo and 120 mg per day Posiphen group, suggesting a drug effect on lowering APP production (Figure 6). Additionally, the single 180 mg per day subject had a low APP production rate and an unusual SILK curve with a very late peak in labeling (about 28 hours), consistent with a drug effect as a previous study with 88 subject (33 amyloid positive) showed a maximum peak in labeling of 24.7 h. However, PK parameters for Posiphen or its metabolites, such as AUC, did not show a correlation with the decrease in APP production rates (Supplementary Figure 1).

**Figure 5.**
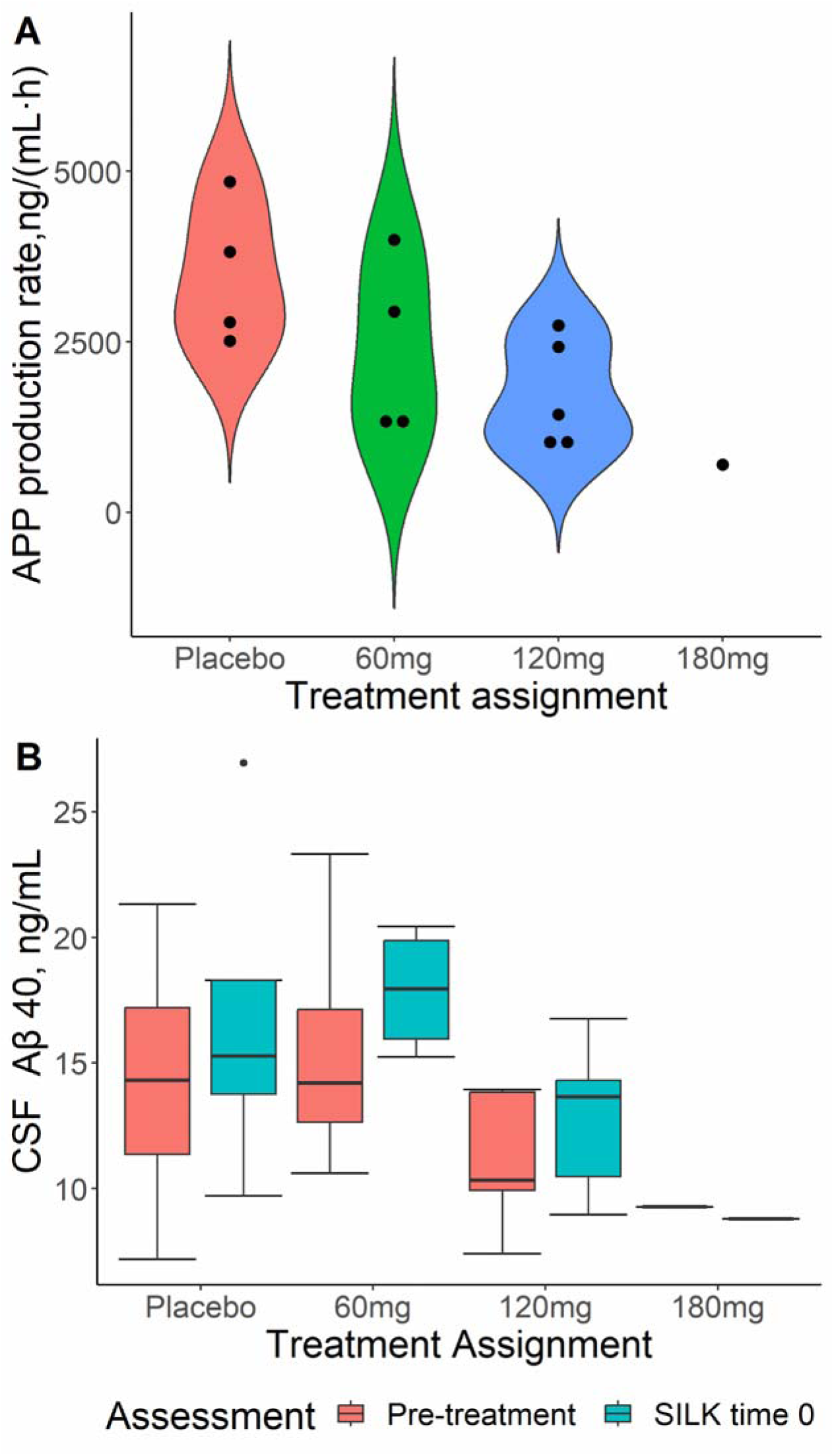
A) APP production rate estimates by treatment arm. B) CSF Aβ40 concentrations at pre-treatment (red) and after 21 days of treatment (cyan).

**Figure 6.**
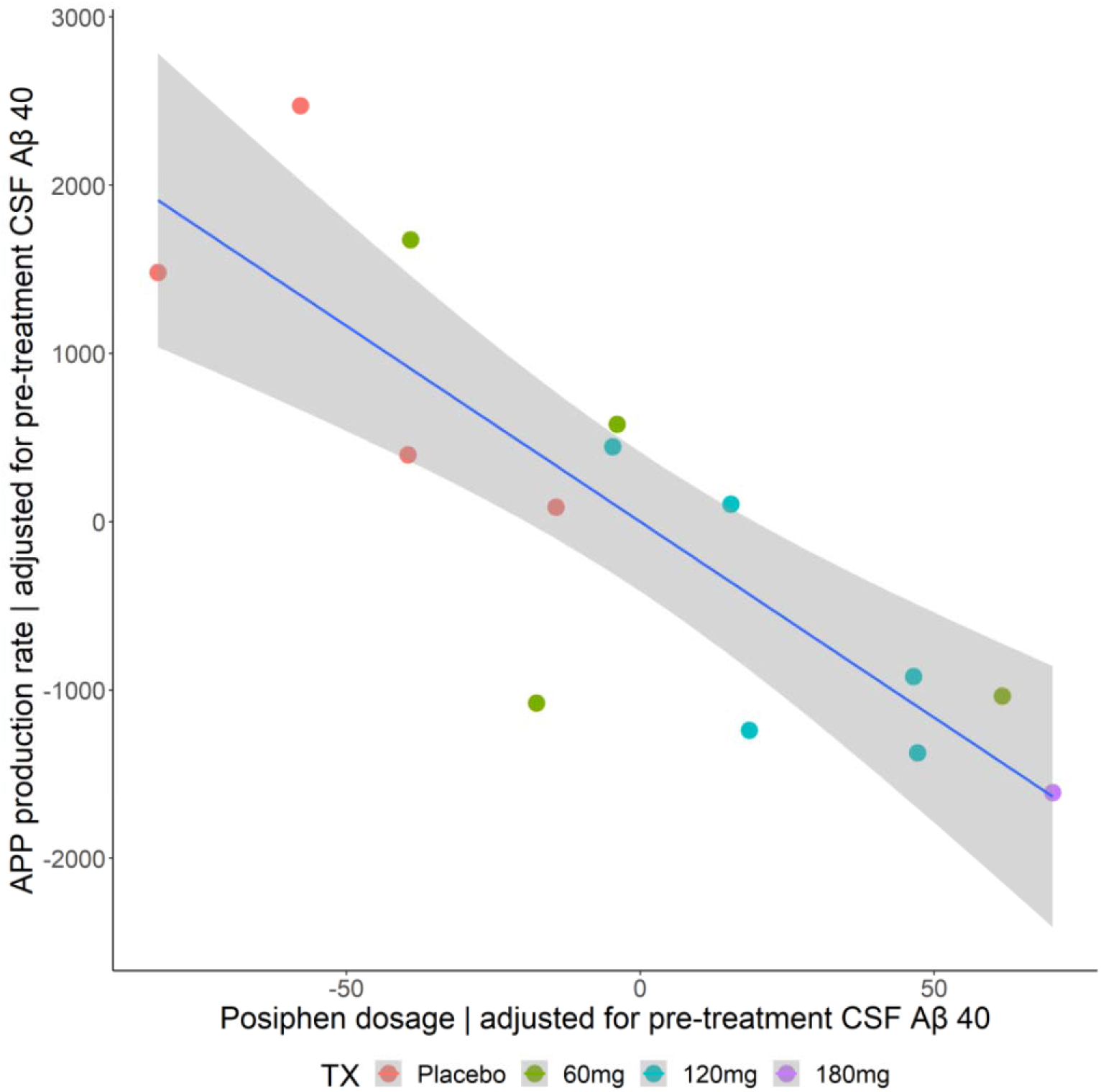
Estimated APP production vs Posiphen dose adjusted for pre-treatment CSF Aβ40.

### Cognition and behavior

There were no significant changes in the ADAS-cog, MMSE or NPI total scores from baseline to the pre-confinement visit in the Posiphen or placebo groups (Supplementary table 1).

### Feasibility of a multi-center SILK study

The quality of sample collection for SILK analysis was a key measure of feasibility and was supported by detailed training of sites and development of protocols and SOPs. Collection of CSF samples was effective (81.75% of CSF samples of 6 mL at 19 time points were obtained from 15 completing participants); matching plasma samples were collected at 100% of these time points. The quality of CSF and plasma samples collected during the confinement visits was sufficient to allow SILK analyses to be carried out on all participants who underwent successful serial CSF sampling. A subject satisfaction survey was completed both following the baseline visit to the trial as well as following the confinement visit. At both visits > 80% of participants rated almost all or most of their expectations having been met (Table 6). Following the confinement visit, features that were best liked about the study included volunteering (31%), raising awareness (25%), interacting with staff (25%) and others (18%). Of features least liked were the time commitment (31%), testing procedures (19%), lack of feedback of results (12%) and others (31%).

**Table 6.**
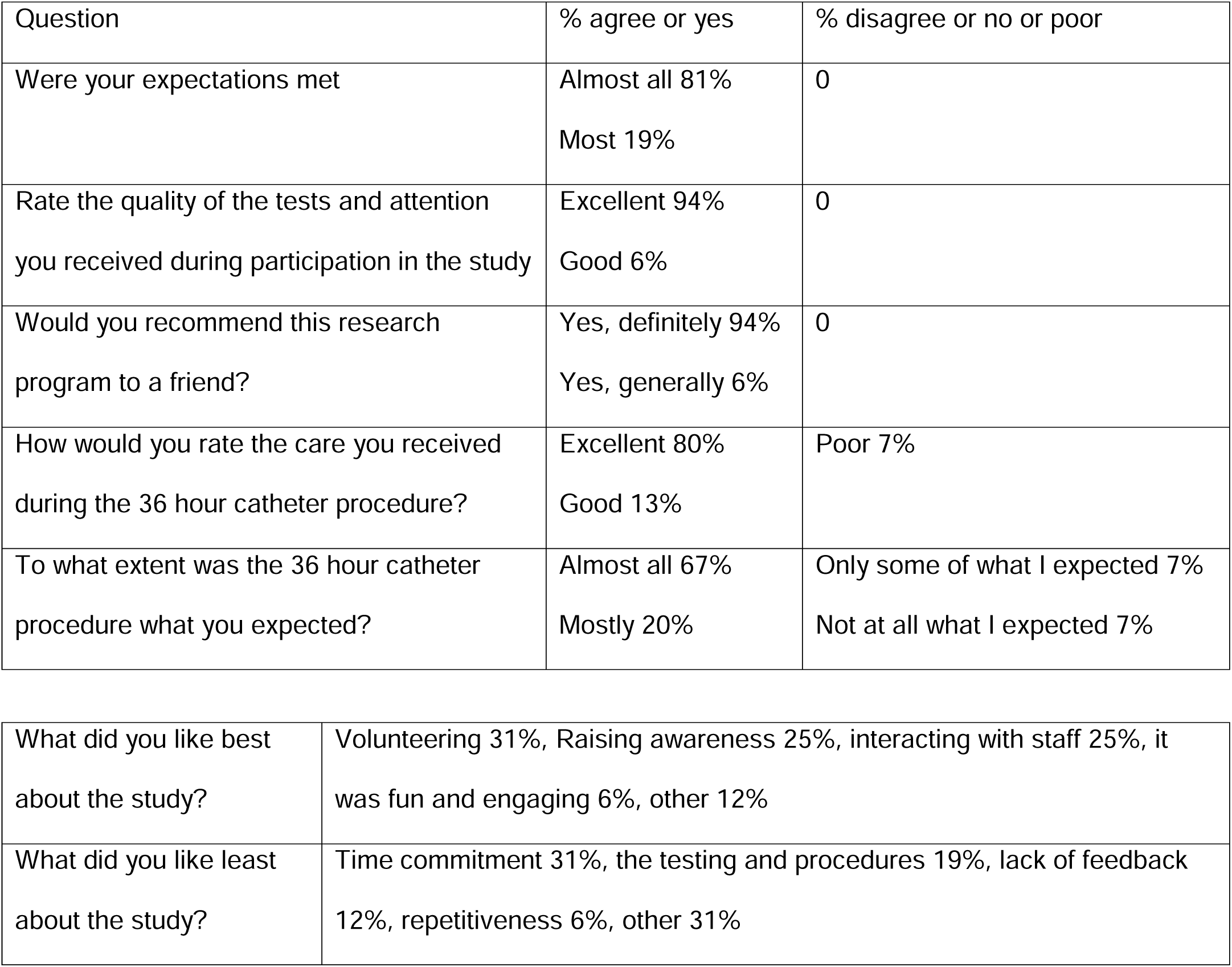
Responses to Study Satisfaction Survey (at confinement visit) (n = 18)

| Question | % agree or yes | % disagree or no or poor |
| --- | --- | --- |
| Were your expectations met | Almost all 81%<br>Most 19% | 0 |
| Rate the quality of the tests and attention you received during participation in the study | Excellent 94%<br>Good 6% | 0 |
| Would you recommend this research program to a friend? | Yes, definitely 94%<br>Yes, generally 6% | 0 |
| How would you rate the care you received during the 36 hour catheter procedure? | Excellent 80%<br>Good 13% | Poor 7% |
| To what extent was the 36 hour catheter procedure what you expected? | Almost all 67%<br>Mostly 20% | Only some of what I expected 7%<br>Not at all what I expected 7% |

|  |  |
| --- | --- |
| What did you like best about the study? | Volunteering 31%, Raising awareness 25%, interacting with staff 25%, it was fun and engaging 6%, other 12% |
| What did you like least about the study? | Time commitment 31%, the testing and procedures 19%, lack of feedback 12%, repetitiveness 6%, other 31% |

## DISCUSSION

Posiphen was safe and well tolerated over 21 days of treatment in Early AD. This augments data from prior studies that have been conducted with varying duration of exposure to Posiphen in healthy volunteers and in people with AD and Parkinson’s Disease.^21,37^ Safety and tolerability did not emerge as concerns among study participants who were also taking acetylcholinesterase inhibitors.

The DISCOVER trial provides evidence that a multicenter SILK study protocol can be successfully implemented with good acceptance by participants. Its feasibility was established with evidence of successful serial two-hourly samples of plasma and CSF over a 36 hour period according to schedule and in a way that adequately supported SILK analyses. Training of site personnel, including a site training visit to Washington University, and the preparation of a video by ADCS Clinical Operations staff showing the steps in setting up a SILK study, were important elements to assist implementation and standardization.

The DISCOVER study was underpowered by virtue of incomplete enrollment due to COVID 19-related delays. In particular, although the 180 mg dose arm yielded interesting data in the single participant who received active drug, we cannot model pK or PD within that dose cohort, nor is sufficient safety data available for that dose. The first phase one study of Posiphen used single ascending doses ranging from 10 to 160 mg/day, and 20 to 60 mg four times per day for 10 days in multiple dose studies, with CSF biomarker determination in a small MCI cohort before and after 10 days of 60 mg four times per day,^20^ Gastrointestinal and other symptoms emerged at the dose of 60 mg 4 times per day and hence 4×60 mg four times a day was determined the no observed adverse effect level. This served as background for our exploration of 60 mg given 1-3 times per day.

The PK data are similar to those previously published, demonstrating a short half-life of Posiphen and its metabolites in plasma and a somewhat longer half-life, with relatively low peak concentration of free drug, in CSF. The data also show similar PK trajectories for Posiphen as for its active metabolites.

Overall PD results showed no difference in the prespecified outcome measure of FSR of Aβ40 by treatment arm. The single 180 mg subject had unusual SILK data, with a possible effect on CSF APP metabolism. By applying an advanced model of APP metabolism to the subjects and data in this trial, we found a suggestive relationship between Posiphen dose and decreased APP production. Longitudinal changes of CSF biomarkers over 21 days were small, but many were in a direction that could support an effect of Posiphen on translational inhibition of APP. In a recently published study of Posiphen in patients with AD and PD, there were small changes in CSF biomarkers over 17 days of treatment at lower doses than we studied.^21^

Posiphen is rapidly metabolized, so it is unclear how long CNS effects on APP production rates might last after drug exposure. The 120 mg per day group received two 60 mg doses at 12 hour intervals, while the 180 mg per day group received three 60 mg doses at 8 hour intervals. Analysis of metabolite kinetics in plasma and CSF did not reveal a higher AUC for the 120 mg per day group versus the 60 mg per day group. This may be due to lack of sensitivity in the measurement, but the lower APP production rates in the 120 mg group from the post hoc model would be expected to be correlated with a higher AUC. The lumbar CSF concentration of Aβ peptides did not decrease significantly after three weeks of Posiphen exposure, although variability of these levels and the small number of participants limits power to detect this change. It is possible that the Aβ CSF concentration was buffered by the presence of plaques in the subjects, but it is unclear why Aβ38 and Aβ40 CSF concentration would be affected by plaques. The trend towards lower CSF concentrations of Aβ40 peptides in the 120 mg and 180 mg participants at study screening was noted and addressed in the model of APP production by controlling for these pre-drug levels. In doing so there was a downward trend with increasing dose. Although the model-derived APP production rate and the screening CSF Aβ40 both decreased with dose, the significant correlation when analyzed together suggests that the decline in model-derived APP production rate was greater than can be explained by natural variation in CSF Aβ40. The reduction in APP production associated with Posiphen is therefore a tentative finding, due to the relatively small numbers of participants, variability and group differences in levels of Aβ40 at entry. Determining APP production rates by SILK before and after drug treatment would provide more precise estimates of changes but presents considerably greater challenges in subject recruitment and retention and overall cost.

A recent study in mice and cognitively normal humans (aged 18-50) using SILK showed that citalopram (as a single dose of 60 mg) decreased Aβ production.^38^ However, a secondary analysis of Alzheimer’s Disease Neuroimaging Initiative data found that people with Mild Cognitive Impairment or AD who were taking SSRI medications did not show differences in amyloid PET burden or in rates of cognitive decline.^39^ In our Posiphen study, more people in the placebo group were taking one or more antidepressant medications than in the treatment groups, and at lower doses (or equivalents) than the 60 mg used in the single dose study. If there was an effect of SSRI or similar medication use on APP or Aβ metabolism among our study participants, it could have made it harder to find an additional benefit due to Posiphen.

A limitation for accepting the tentative findings for APP synthesis and Posiphen in this study was the lack of a decrease in CSF concentrations of sAPPα and sAPPβ on serial lumbar punctures. These have been noted to decline in previous studies of Posiphen; however, levels of these products of APP in CSF have substantial variability and our study was not powered to detect a change. The fact that the single participant receiving the drug three times per day showed anomalous SILK kinetics in the expected direction (exceptional when compared to 88 prior SILK curves) is noteworthy, but it is difficult to make firm conclusions from this single set of data points.

## CONCLUSIONS

The proposed mechanism of action of Posiphen to inhibit APP translation did not result in treatment-dependent reduction of FSR of Aβ40 in people with Early AD. However, the advanced SILK modeling of APP kinetics provided tentative support for this mechanism. Overall, this study provided additional safety, PK and PD data to help to advance Posiphen for future clinical trials, although the best dosing regimen remains unclear. The multisite methods developed here may also be applicable to the study of other drugs that alter Aβ kinetics.

## ABBREVIATIONS

Aβ: amyloid beta protein
AD: Alzheimer’s Disease
ADAScog: Alzheimer’s Disease Assessment scale-cognitive
ADCS: Alzheimer’s Disease Cooperative Study
APP: Amyloid Protein Precursor
AUC: Area Under the Curve
Cmax: maximal concentration
CSF: cerebrospinal fluid
DSMB: Data Safety Monitoring Board
EKG: electrocardiogram
FSR: Fractional Synthesis Rate
ISF: interstitial fluid
NPI: Neuropsychiatric Inventory
PD: pharmacodynamics
PK: pharmacokinetics
sAPPα: secreted APP alpha
sAPPβ: secreted APP beta
SILK: Stable Isotope Labeling Kinetics

## DECLARATIONS

### Ethics approval

the study was approved by the UCSD Human Research Protection Program, operating under Federalwide Assurance number, FWA00004495. All participants provided informed consent.

### Consent for publication

not applicable

### Data Availability

Data and biological samples from this study are available to qualified investigators upon request to the ADCS. Requests will be reviewed by the ADCS Data and Sample Sharing Committee. The code for the detailed compartmental modeling of APP metabolism is available from Dr Elbert.

### Competing interests

MM is an employee of Annovis Bio, TW is an employee of C2N Diagnostics, RB is a cofounder of C2N Diagnostics

### Funding

Grant U19 AG010483 from the National Institute on Aging. Study drug and placebo were provided by Annovis.

### Authors’ Contributions

DG obtained funding, designed the study, obtained participant data, helped with analysis and interpretation and drafted the manuscript. MF helped to design the study, obtained participant data, helped with analysis and interpretation and helped revise the manuscript. BPL obtained participant data, helped with analysis and interpretation and helped revise the manuscript. LSH obtained participant data, helped with analysis and interpretation and helped revise the manuscript. DE helped with analysis and interpretation and helped revise the manuscript. RB helped with analysis and interpretation and helped revise the manuscript. JM helped with analysis and interpretation and helped revise the manuscript. RT helped with analysis and interpretation and, helped revise the manuscript. RAR helped with sample storage and analysis and helped revise the manuscript JP helped with analysis and interpretation and helped revise the manuscript. VA helped with analysis and interpretation and helped revise the manuscript. AB helped with study implementation and conduct and helped revise the manuscript. TW helped with study implementation and helped revise the manuscript. MM helped with study implementation and helped revise the manuscript. HHF helped with study design, implementation and oversight, helped analyze and interpret data and helped revise the manuscript. All authors have read and approved the final manuscript.

## Acknowledgments

We acknowledge the efforts of research staff and Faculty at the following sites: UC San Diego: Dan Szpak, RN (Study Coordinator); Mark Wallace, MD (Anesthesiologist); Indiana University: Gena Antonopoulos, RN and Brooke Boersma, RN (Study Coordinators); Washington University: Cristina Toedebusch and Tiara Redrick (Study Coordinators) and Robert Swarm, MD (Anesthesiologist); Columbia University : Evelyn D. Dominguez, MD (Study Coordinator) and Wendy P. Gonzalez, NP (Nurse Practitioner); Abhay Moghekar, MD (Johns Hopkins Hospital). We also acknowledge the support of Kevin Yarasheski and Philip Verghese (C2N Diagnostics). We acknowledge support from Annovis by providing study drug and serving as study sponsor.

## Notes

### Competing Interest Statement

The authors have declared no competing interest.

### Clinical Trial

NCT02925650

### Author Declarations

UCSD Human Research Protections Program

