## Supplementary data for "A multicenter, randomized, double-blind, placebo-controlled ascending dose study to evaluate the safety, tolerability, pharmacokinetics (PK) and pharmacodynamic (PD) effects of Posiphen in subjects with Early Alzheimer’s Disease"

**Supplementary Table 1. Changes in ADAS-cog12, MMSE and NPI in relation to treatment with Posiphen vs placebo**

|  | Group | Baseline | Pre-confinement | Mean change | Treatment difference (Posiphen – placebo) |
| --- | --- | --- | --- | --- | --- |
| **ADAS-Cog12** | Placebo (n=7) | 24.86 ± 9.62 | 23.29 ± 9.95 | -1.57 ± 5.19 | 3.00 ± 2.94 p = 0.32 |
|  | Posiphen (n = 11) | 25.73 ± 8.76 | 25.18 ± 5.33 | -0.55 ± 6.04 |  |
| **MMSE** | Placebo (n=7) | 24 ± 4.4 | 25.14 ± 4.78 | 1.14 ± 2.12 | -1.6 ± 1.28  p= 0.23 |
|  | Posiphen (n = 11) | 24.36 ± 2.8 | 24.18 ± 2.52 | -0.18 ± 2.09 |  |
| **NPI** | Placebo (n=7) | 16.29 ± 14.94 | 12.43 ± 10.24 | -3.86 ± 8.09 | 7.10 ± 4.07 p = 0.11 |
|  | Posiphen (n = 11) | 8.36 ± 9.52 | -0.45 ± 7.01 | 0.107 |  |

mITT analyses were performed. In separate analyses, there was no significant effect of dose arm on change of ADAS-cog12, MMSE or NPI.

**Supplementary Table 2. Changes in ADAS-cog12, MMSE and NPI for each dose group compared to placebo**

| Contrast | ADAS-cog | | MMSE | | NPI | |
| --- | --- | --- | --- | --- | --- | --- |
|  | t | p | t | p | t | p |
| 60 mg – placebo | 0.42 | 0.68 | -3.32 | 0.006 | 0.68 | 0.51 |
| 120 mg - placebo | 1.1 | 0.30 | 1.2 | 0.24 | 2.73 | 0.02 |
| 180 mg - placebo | 029 | 0.77 | -1.9 | 0.85 | 0.12 | 0.91 |

Multiple linear regression, including age, sex, APOE genotype, baseline cognitive score and arm in the model.

**Supplementary Figure 1. Correlations between estimates of APP production rates and pharmacokinetic parameters.**

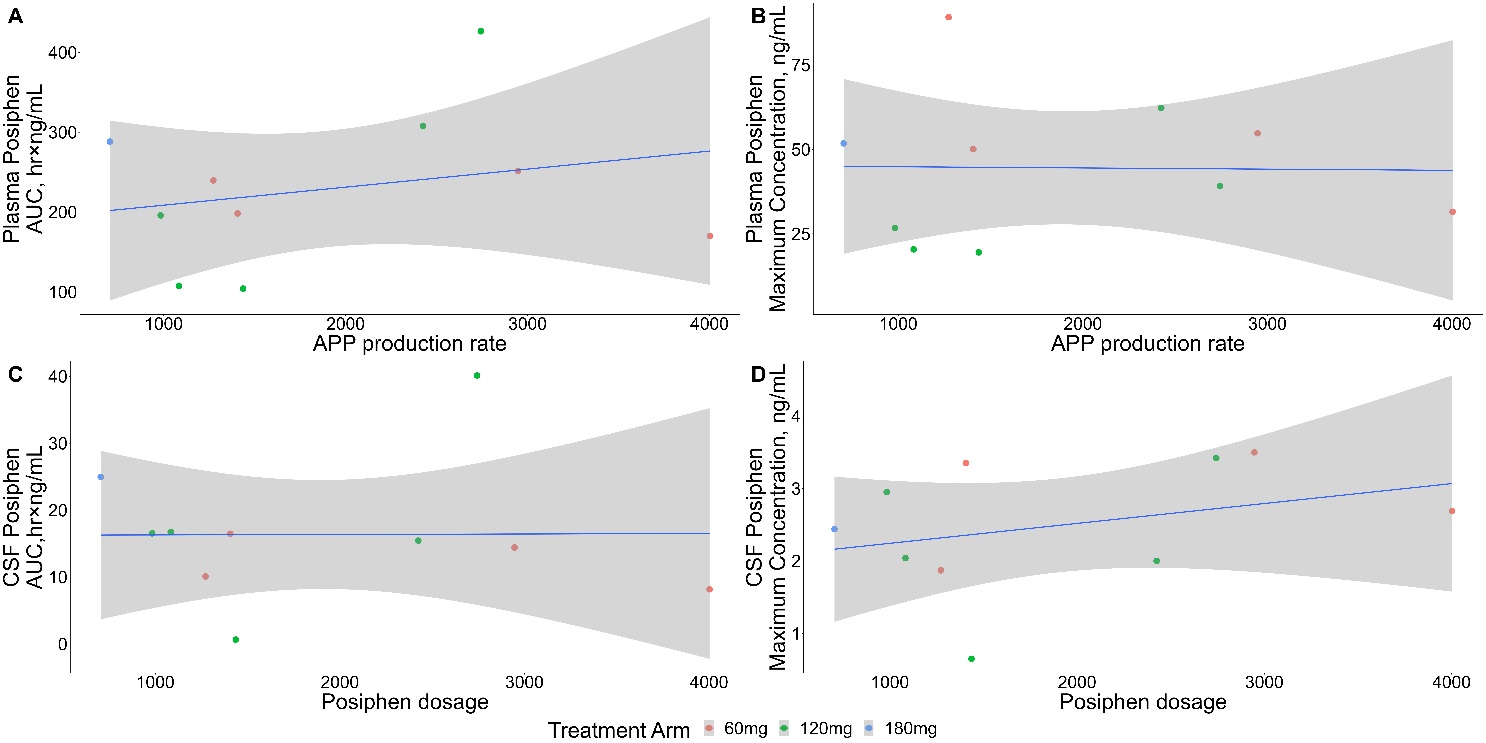
